# Alternative dosing regimens of GLP-1 receptor agonists may reduce costs and maintain weight loss efficacy

**DOI:** 10.1101/2024.11.27.24318093

**Authors:** Anıl Cengiz, Calvin C. Wu, Sean D. Lawley

## Abstract

**Aims:** To discover alternative dosing regimens of incretin mimetics that simultaneously reduce costs and maintain weight loss efficacy. As a secondary objective, we used our results to explore how allocating a limited incretin mimetics budget could affect public health on a national scale.

**Materials and Methods:** We used mathematical modeling and simulation of semaglutide and tirzepatide. For semaglutide, we used a recent pharmacokinetic (PK) and pharmacodynamic (PD) model. For tirzepatide, we used a recent PK model and modeled PD by reparameterizing the semaglutide PD model to fit tirzepatide clinical data.

**Results:** Reducing dose frequency does not commensurately reduce weight loss. For example, merely switching from one dose per week (q1wk) to one dose every two weeks (q2wk) maintains roughly 75% of the weight loss. Furthermore, if the decrease in dose frequency involves an appropriate increase in dose size, then approximately 100% of the weight loss is maintained. In addition, we compared offering incretin mimetics to (1) a fraction of obese US adults with q1wk dosing versus (2) twice as many obese US adults with q2wk dosing. Though scenarios (1) and (2) require the same budget, our analysis suggests that (2) reduces national obesity and mortality to a much greater degree.

**Conclusion:** Our study highlights the potential utility of alternative dosing regimens of incretin mimetics. Compared to standard once-weekly dosing, costs can be halved and weight loss maintained. These cost-saving results have implications for patients, physicians, insurers, and governments.

## 1 Introduction

Obesity is the most prevalent chronic disease worldwide [1] and drives significant morbidity and mortality. Obesity increases the risk of a variety of other chronic diseases, including type 2 diabetes, heart disease, and certain cancers [2, 3]. In the United States (US), approximately 71% of adults are overweight (defined as having a Body Mass Index (BMI) in kg*/*m^2^ greater than or equal to 25), including 41% who are obese (BMI *≥* 30) and 8% who are severely obese (BMI *≥* 40) [4]. The financial costs of excess adiposity are massive, as it is estimated that obesity results in over $170 billion in healthcare spending in the US every year [5].

We are currently in the midst of a revolution in weight loss interventions. Indeed, while both lifestyle and older pharmaceutical approaches have demonstrated limited efficacy [6, 7], glucagon-like peptide-1 (GLP-1) and dual GLP-1/gastric inhibitory polypeptide (GLP-1/GIP) receptor agonists have recently been shown to yield unprecedented levels of weight loss [8, 9]. For example, the incretin mimetics semaglutide and tirzepatide have been shown via meta-analyses of randomized controlled trials to induce placebo-adjusted average body weight losses of 15.0% and 19.2%, respectively [10].

Despite their demonstrated efficacy and safety, patient access to incretin mimetics has been severely limited for two primary reasons. The first reason is supply shortages which have left many patients unable to fill their prescriptions. Indeed, supply shortages were identified as a potential reason for the rather low persistence and adherence rates reported for semaglutide [11].

The second reason, which is a more daunting and likely persistent problem, is the very high cost of these medications [12]. Often in excess of $1,000 per month without insurance, these medications are simply not financially feasible for many individuals who stand to benefit from them. At the national scale, these high costs prompted a US Senate Committee hearing [13] amidst fears that this new generation of anti-obesity medications could bankrupt the US healthcare system [14]. In fact, providing each eligible US adult with a GLP-1 would roughly double the total prescription drug spending in the US [15].

In this paper, we use mathematical modeling and simulation to investigate the weight loss efficacy of GLP-1 receptor agonists under alternative dosing regimens. We seek dosing regimens which both (i) reduce cost and (ii) maintain high efficacy compared to the standard once per week dose. The specific incretin mimetics that we investigate are semaglutide (brand name Wegovy for weight loss and Ozempic and Rybelsus for type 2 diabetes) and tirzepatide (brand name Zepbound for weight loss and Mounjaro for type 2 diabetes). We further investigate how alternative dosing regimens could affect public health on the national scale.

## 2 Methods

We now briefly summarize our methods. The full details are presented in the Supplementary Appendix. For semaglutide, we use the pharmacokinetic (PK) and pharmacodynamic (PD) model proposed and validated by Strathe et al. [16]. The PK model is a single compartment model with linear absorption and elimination. The PD model (i.e. exposure-response model) is a semi-mechanistic, non-linear model. The PKPD model recapitulates longitudinal weight loss data from three randomized, double-blind, controlled trials of subcutaneous semaglutide [9, 17, 18]. For tirzepatide, we use the two compartment, linear PK model proposed and validated by the US Food and Drug Administration [19]. For tirzepatide PD, we reparameterize the semaglutide PD model to fit the longitudinal weight loss data from the double-blind, randomized, controlled trial of Jastreboff et al. [8].

To explore the effects of alternative dosing regimens on the national scale, we estimate how the BMI distribution of adults in the US would change if different proportions of obese US adults lost various proportions of body weight. We then translate these changes in BMI distribution into estimates of changes in mortality using the BMI-dependent mortality rates reported by Pandey et al. [4].

## 3 Results

### 3.1 Reparameterized semaglutide model fits tirzepatide weight loss data

In Figure 1, we compare the tirzepatide PD model to the tirzepatide longitudinal weight loss data from Jastreboff et al. [8]. This plot demonstrates excellent agreement between the model and the clinical trial data (the data is from Figure 1B in [8]). Indeed, the model values are all within 1% of the data for the relative change in body weight across placebo, 5 mg, 10 mg, and 15 mg doses of tirzepatide measured at 11 time points over the 72 week study. In Figure 1, tirzepatide was initiated at a dose of 2.5 mg once weekly (except placebo) and was increased by 2.5 mg every 4 weeks up to a maintenance dose of either 5 mg, 10 mg, or 15 mg. To our knowledge, this is the first PKPD model to recapitulate the tirzepatide weight loss data in [8].

**Figure 1:**
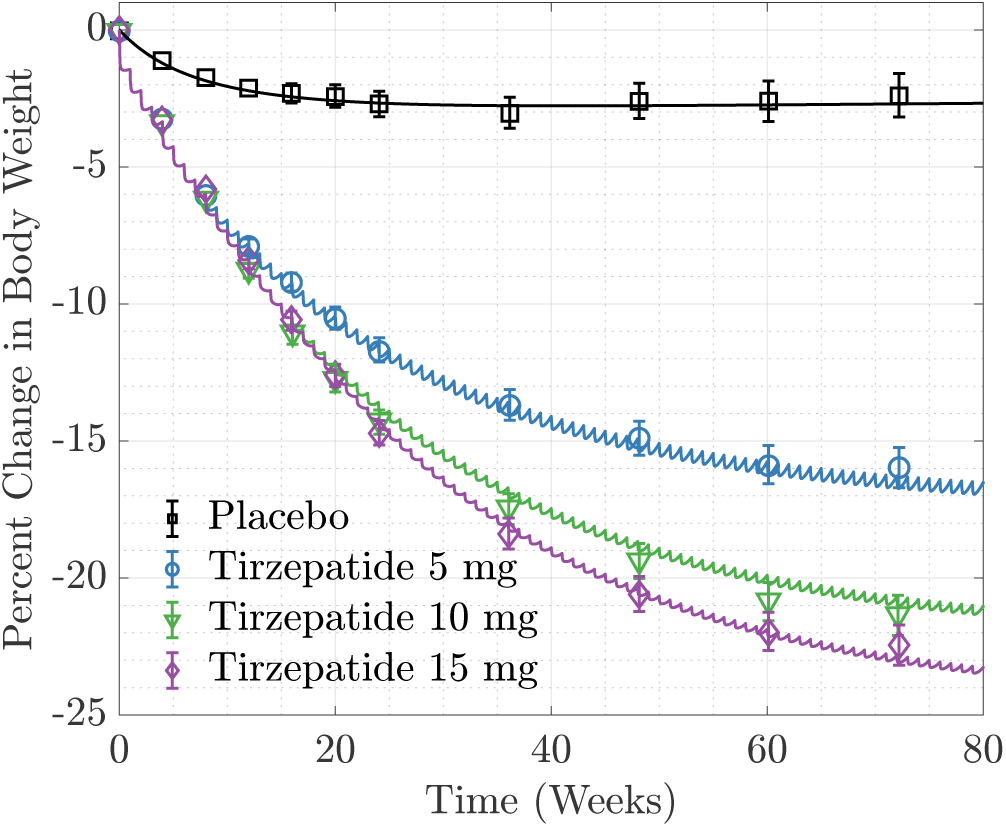
Reparameterized semaglutide PD model fits tirzepatide weight loss data. Solid curves are the tirzepatide PD model. Markers are clinical trial data from Jastreboff et al. [8], where the error bars around each data point indicate 95% confidence intervals.

We obtained the tirzepatide PD model by reparameterizing the semaglutide PD model of Strathe et al. [16] to fit the data from Jastreboff et al. [8]. Since these PD models of tirzepatide and semaglutide are structurally identical, we can directly compare their parameter values (see the Supplementary Appendix). Interestingly, the tirzepatide PD parameters estimated in this work are quite similar to the semaglutide PD parameters estimated by Strathe et al. [16]. The similarity in parameter values for tirzepatide and semaglutide is not surprising given the similar efficacies observed in clinical trials [8, 9].

### 3.2 Less frequent dosing of semaglutide

Though dose sizes vary between patients, both semaglutide and tirzepatide are typically administered once per week (q1wk). Indeed, once-weekly dosing is the dosing frequency studied in clinical trials [8, 9, 17, 20–22] (Ref. [18] studied once-daily semaglutide). We now use the mathematical models of semaglutide and tirzepatide to investigate the weight loss efficacy of alternative dosing regimens. Figure 2 predicts the efficacy of semaglutide under less frequent dosing. In Figure 2A, we plot the steady state percent change in body weight as a function of the dosing interval (time between doses) for the standard 2.4 mg dose of semaglutide. The most salient feature of Figure 2A is that increasing the dosing interval (i.e. decreasing the dosing frequency) does not commensurately decrease the weight loss efficacy of semaglutide. For instance, for the standard once-weekly (q1wk) dosing (i.e. a dosing interval of 7 days), the model predicts a steady state body weight reduction of 17%, which is in good agreement with clinical data for q1wk dosing [10,16]. For a dosing interval of 14 days (q2wk), the model predicts a steady state body weight reduction of 12%. Therefore, despite the fact that decreasing the dosing frequency from q1wk to q2wk decreases the total amount of drug taken over time by one half, the model predicts that patients retain 72% of their weight loss compared to q1wk. Furthermore, the model predicts that nearly 50% of weight loss is retained when comparing once-weekly to only once-monthly dosing (i.e. comparing a 7 versus 28 day dosing interval in Figure 2A), though clinical validation is especially warranted for this prediction.

**Figure 2:**
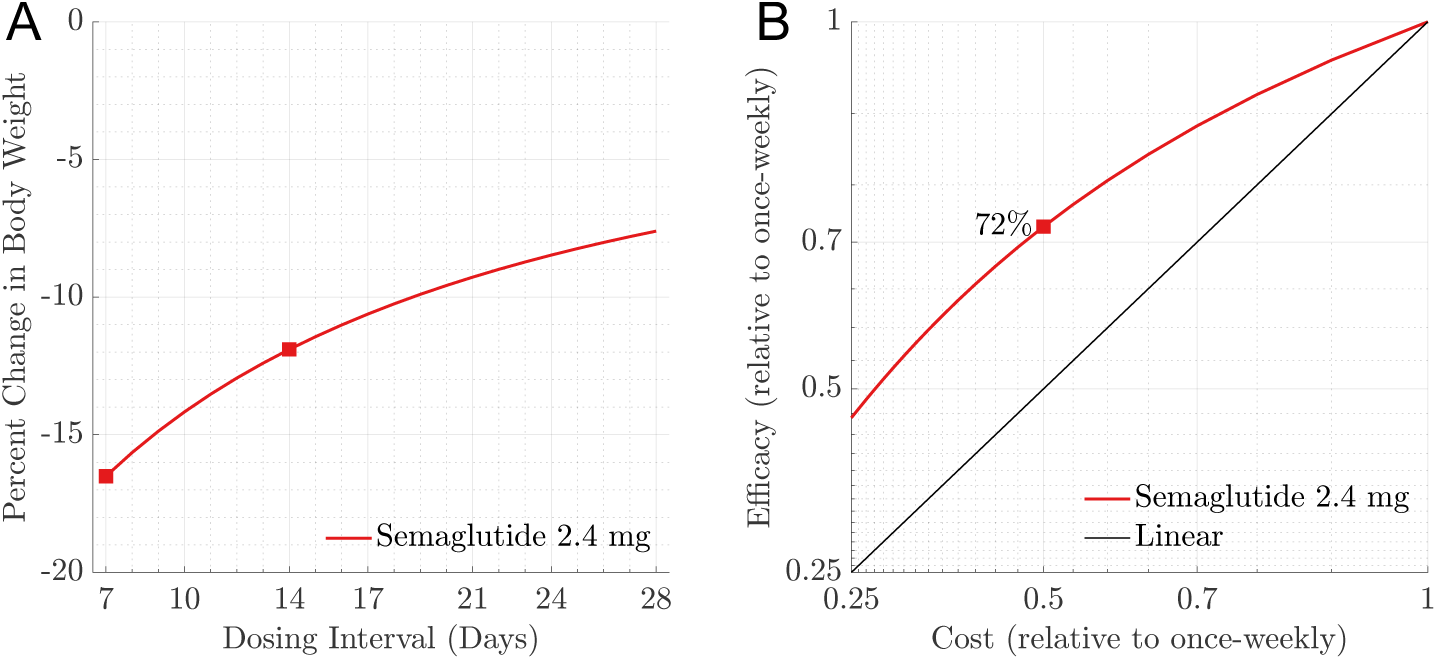
Increasing the time between doses of semaglutide does not commensurately decrease weight loss. Panel A: Steady-state percent change in body weight as a function of the dosing interval (time between doses). The markers highlight weight loss for once weekly dosing (q1wk) versus once every other week dosing (q2wk). Panel B: How the steady-state weight loss (efficacy) and cost decrease when doses are taken less frequently than once-weekly. The marker highlights that switching from q1wk to q2wk retains 72% and reduces cost by 50%.

These points are illustrated in Figure 2B, where we plot the weight loss efficacy relative to q1wk against the cost relative to q1wk. This plot assumes that cost is proportional to the number of doses, and the reduction in cost comes from reducing the dosing frequency. Hence, for a fixed dose size of 2.4 mg of semaglutide, reducing dose frequency can reduce costs and maintain strong efficacy.

### 3.3 Alternative dosing of tirzepatide

Figure 3 predicts the efficacy of tirzepatide under alternative dosing regimens (decreasing dose frequency and potentially increasing dose size). In Figure 3A, we plot the steady state percent change in body weight as a function of the dosing interval (time between doses) for 5 mg, 10 mg, and 15 mg doses of tirzepatide. Analogous to Figure 2A, decreasing the dose frequency does not commensurately decrease the weight loss efficacy of tirzepatide. For instance, for the standard q1wk dosing, the model predicts steady state body weight reductions of 17%, 21%, and 23% for tirzepatide at 5 mg, 10 mg, and 15 mg, respectively (which is in good agreement with clinical data [8, 10]). For q2wk dosing, the model predicts steady state body weight changes of 12%, 16%, and 18% for tirzepatide at 5 mg, 10 mg, and 15 mg, respectively. Hence, similar to semaglutide, the model predicts that patients retain roughly 75% of their weight loss when merely switching from q1wk to q2wk dosing.

**Figure 3:**
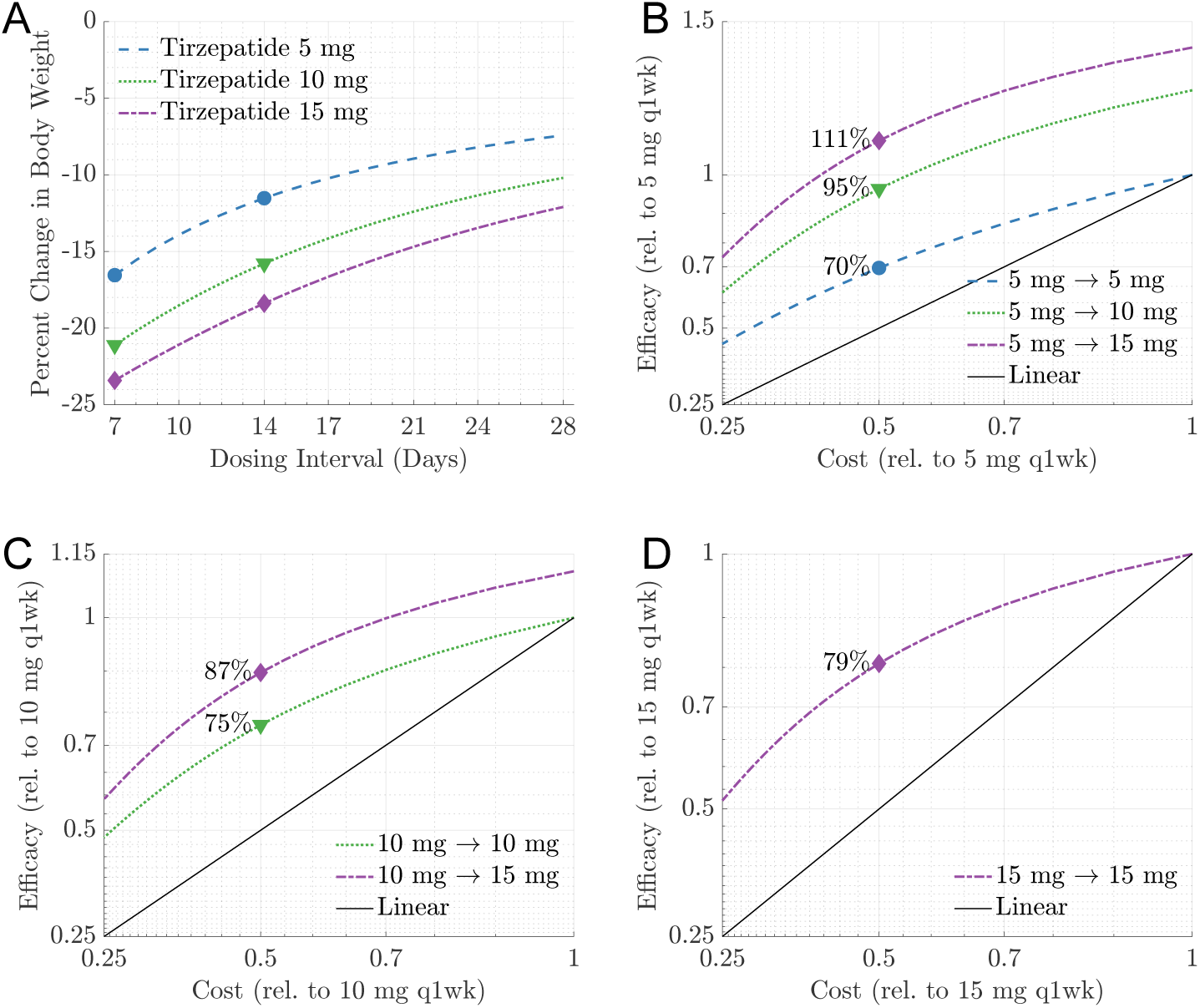
Alternative dosing regimens of tirzepatide decrease costs and maintain efficacy. Panel A: Steady-state percent change in body weight as a function of the dosing interval (time between doses). The markers highlight weight loss for once weekly dosing (q1wk) versus once every other week dosing (q2wk). Panel B: How a patient currently on 5 mg q1wk can decrease costs and maintain (or improve) efficacy. The markers indicate that switching to q2wk decreases costs by 50% and maintains (i) 70% of their weight loss if the dose size is kept at 5 mg, (ii) 95% of their weight loss if the dose size is doubled to 10 mg, and (iii) 111% of their weight loss if the dose size is tripled to 15 mg. Analogously, Panels C and D concern patients currently on 10 mg q1wk and 15 mg q1wk, respectively.

Tirzepatide is commercially available in dose sizes varying from 2.5 mg to 15 mg in increments of 2.5 mg. We thus investigate alternative dosing regimens that vary the dose frequency and the dose size. For instance, Figure 2A predicts that 5 mg taken every 7 days yields approximately the same steady state weight loss as 10 mg taken every 14 days. Similarly, Figure 2A predicts that 10 mg taken every 7 days yields approximately the same steady state weight loss as 15 mg taken every 10 days.

Importantly, the current standard pricing structure of tirzepatide charges patients per dose (injection), regardless of the dose size [23] (the same is true for semaglutide [24]). For example, a 2.5 mg dose is the same price as a 15 mg dose. Therefore, increasing the dosing interval and appropriately increasing the dose size can significantly reduce costs with essentially zero difference in efficacy. The cost saving potential of such alternative dosing regimens is illustrated in Figure 2B-D.

Figure 2B concerns a patient who currently takes 5 mg of tirzepatide at the standard once-weekly dosing. If this patient continues to take doses of size 5 mg, then the blue dashed curve shows how the efficacy decreases as the cost decreases by decreasing the dose frequency. For instance, switching from q1wk to q2wk but keeping a 5 mg dose size decreases cost by 50% and maintains 70% of weight loss. If the patient increases the dose size to 10 mg in addition to decreasing the dose frequency, then the green dotted curve shows the resulting efficacy versus cost relationship. Notice that switching from 5 mg q1wk to 10 mg q2wk decreases cost by 50% and maintains 95% of weight loss. The purple curve describes switching from 5 mg to 15 mg and predicts that decreasing the dose frequency from q1wk to q2wk yields 111% of weight loss (i.e. the cost is halved and the weight loss is increased).

Figure 2C is analogous to Figure 2B, but concerns a patient who currently takes 10 mg of tirzepatide at the standard once-weekly dosing. If this patient continues a 10 mg dose but switches to q2wk, then they reduce cost by 50% and maintain 75% of their weight loss. If this patient increases the dose size to 15 mg (purple curve), then (a) increasing the dosing interval to 10 days reduces cost by 30% and maintains identical weight loss and (b) increasing the dosing interval to 14 days reduces cost by 50% and maintains 87% weight loss. Figure 2D concerns a patient on 15 mg q1wk and shows that they can cut their costs in half and retain 79% of their weight loss.

### 3.4 Exploring national implications

The high cost of incretin mimetics severely limits access to these life-saving drugs. Indeed, it was recently estimated that increasing access to incretin mimetics could save at least tens of thousands to perhaps over one hundred thousand lives in the US annually [4]. We now briefly explore the implications of alternative dosing regimens on the national scale.

For simplicity, we focus on semaglutide. Consider a total national amount of semaglutide that allows less than half of obese US adults to take 2.4 mg at the standard once weekly dosing frequency. For this same total amount of semaglutide, twice as many US adults could be on semaglutide if all patients took one dose every other week rather than one dose every week.

In Figure 4A-C, we plot how the percentage of obese US adults could decrease if a fixed annual supply of semaglutide was administered to either (1) a proportion of obese US adults at q1wk dosing (green solid curves) or (2) twice as many obese US adults at q2wk dosing (orange dotted curves). Based on the results in Figure 2, we assume that q1wk dosing yields 17% steady-state weight loss, whereas q2wk dosing yields 12% steady-state weight loss. Though scenario (1) results in more weight loss for each individual on semaglutide, scenario (2) decreases national obesity rates to a much greater degree since twice as many individuals can be treated with q2wk dosing compared to q1wk. For instance, 3.3 billion mg of semaglutide per year would be required for 25% of obese US adults to take the standard 2.4 mg dose q1wk. However, this same amount could supply 50% of obese US adults with a 2.4 mg dose q2wk. Similarly, 6.6 billion mg per year could supply (1) 50% of obese US adults with 2.4 mg q1wk or (2) 100% of obese US adults with 2.4 mg q2wk. The predicted effects on the national BMI distribution for these specific values are indicated in Figure 4 with the circle and square markers. The results are also displayed in Table 1 for convenience.

**Figure 4:**
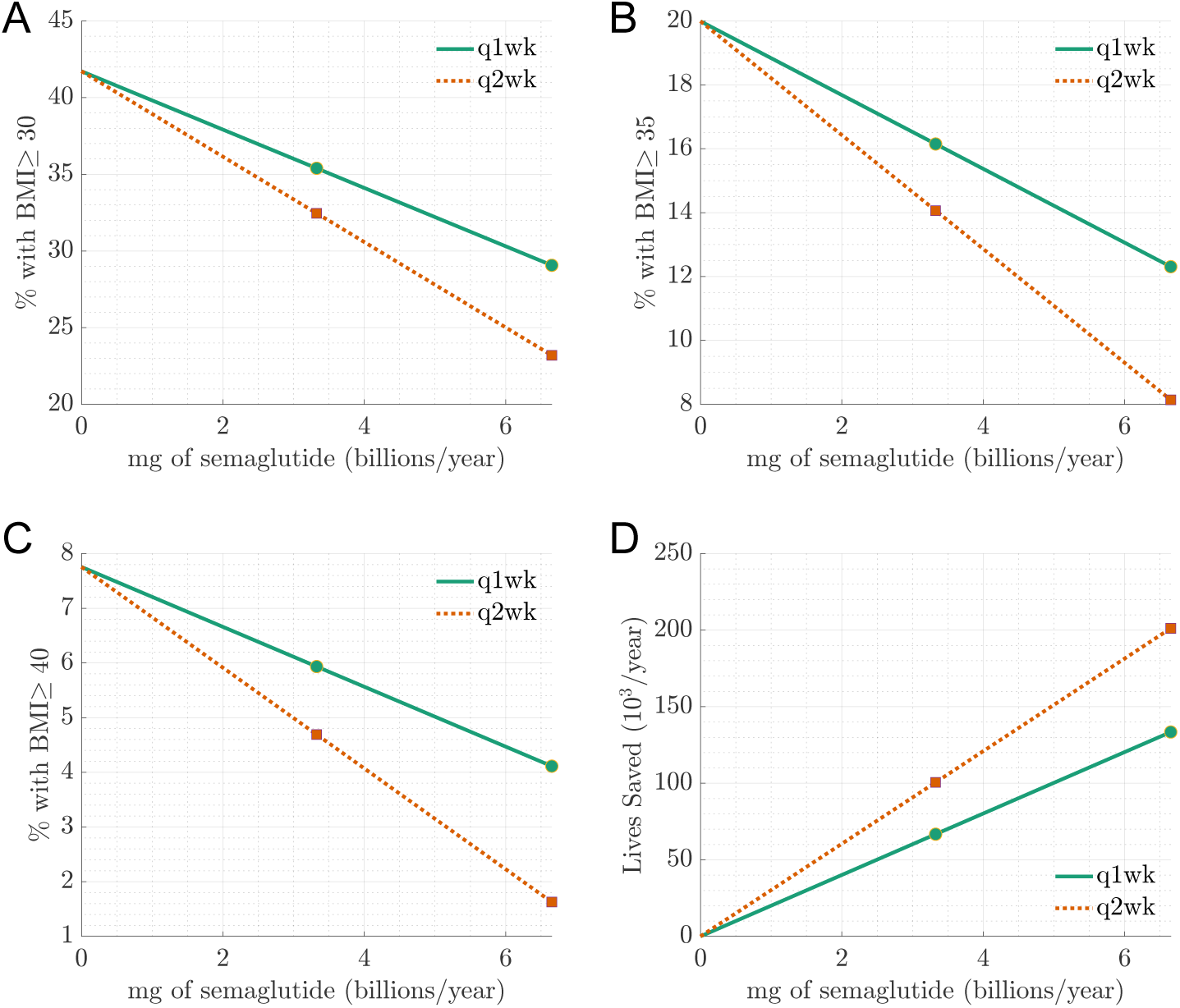
Exploring public health implications of less frequent dosing at the national scale. Panel A estimates how the percentage of obese US adults (BMI*≥* 30) would decrease if a national supply of semaglutide was administered either (1) to some number of obese US adults at q1wk dosing or (2) to twice as many obese US at q2wk dosing. Panels B and C are analogous to Panel A but show the percentage of US adults with Class II Obesity (BMI*≥* 35) and Class III Obesity (BMI*≥* 40), respectively. Panel D uses the BMI distribution predictions to estimate the number of lives saved in scenario (1) versus scenario (2). The markers at 3.3 billion mg per year indicate the amount required for a 2.4 mg dose to be taken by either (1) 25% of obese US adults q1wk or (2) 50% of obese US adults q2wk. Similarly, the markers at 6.7 billion mg per year indicate the amount required for a 2.4 mg dose taken by either (1) 50% of obese US adults q1wk or (2) 100% of obese US adults q2wk.

**Table 1:** US adults by obesity category. The first row is the current BMI distribution. The second row estimates the BMI distribution if 25% of obese US adults take semaglutide once per week (q1wk). The third row estimates the BMI distribution if this same total amount of semaglutide was distributed to 50% of obese US adults with a dose every other week (q2wk). The final two rows are for q1wk dosing to 50% of obese US adults (fourth row) and q2wk dosing to 100% of obese US adults (fifth row). The number of lives saved in the final column is compared to the current BMI distribution and is per year in the US.

| | BMI $\geq 30$ | BMI $\geq 35$ | BMI $\geq 40$ | Lives saved |
| --- | --- | --- | --- | --- |
| Current | 41% | 19% | 8% | - |
| q1wk, 3.3bn mg/year | 35% | 16% | 6% | $0.7 \times 10^5$ |
| q2wk, 3.3bn mg/year | 32% | 14% | 5% | $1.0 \times 10^5$ |
| q1wk, 6.7bn mg/year | 29% | 12% | 4% | $1.3 \times 10^5$ |
| q2wk, 6.7bn mg/year | 23% | 8% | 2% | $2.0 \times 10^5$ |

Following a similar approach as Pandey et al. [4], we use the predicted BMI distribution and BMI-dependent mortality rates to compare the number of lives that could be saved in scenario (1) versus scenario (2). These comparisons are plotted in Figure 4D and are given in the final column of Table 1. Though the quantitative estimates depend on the national semaglutide supply, this analysis predicts that q2wk dosing could save roughly 50% more lives than q1wk dosing. To summarize, owing to the nonlinear relationship between efficacy and dosing frequency/cost predicted in Figure 2, less frequent dosing of incretin mimetics may offer significant population-level public health gains for a given economic cost. We emphasize that the curves in Figure 4 and the numerical values in Table 1 are not precise estimates as they result from very simple calculations (detailed in the Supplementary Appendix). Indeed, weight loss from incretin mimetics varies between patients, specific drugs, and dose sizes [8,9] and depends on persistence and adherence which vary significantly between patients [11, 25] (and an estimated 11% of overweight or obese US adults are strongly opposed to taking weight-loss drugs [26]). In contrast, Figure 4 and Table 1 simply assume a blanket 17% weight loss for one dose per week and 12% weight loss for one dose every other week. Further, these calculations neglect the small percentage of obese US adults currently taking incretin mimetics (recent polling indicates roughly 6% of all US adults are currently on a GLP-1 drug [26]). Nevertheless, these calculations highlight the potential benefits of less frequent dosing given the obesity crisis and current economic realities which severely limit access.

## 4 Discussion

In this paper, we used mathematical modeling and simulation to study how alternative dosing regimens affects the weight loss efficacy of incretin mimetics. We used an existing PKPD model of semaglutide [16] and an existing PK model of tirzepatide [19]. Further, we obtained a new tirzepatide PD model by reparameterizing the semaglutide PD model [9] to fit the tirzepatide clinical trial data reported by Jastreboff et al. [8].

Using these PKPD models, we found that reducing dose frequency does not proportionately reduce weight loss. We proposed alternative dosing regimens that substantially reduce costs and maintain strong efficacy. Indeed, in some scenarios, costs can be halved and weight loss can be maintained at levels which are essentially identical to that obtained under standard dosing regimens. Furthermore, we predicted that less frequent dosing may offer significant public health benefits in terms of reducing national obesity and mortality rates. Hence, in view of the obesity epidemic and the current economic burden of incretin mimetics, alternative dosing regimens may offer significant value both to individual patients and at the broader population level.

Since it was based on mathematical modeling and simulation, our predictions of the weight loss efficacy of alternative dosing regimens requires empirical validation. However, our predictions are supported by the clinical experience of the second author (detailed in the recent case report [27]). In fact, less frequent dosing has been recommended as a strategy to maintain weight loss [15]. Furthermore, our predicted nonlinear relationship between dose frequency and weight loss could be anticipated from clinical data. Indeed, a 100% increase in the weekly tirzepatide dose from 5 mg to 10 mg increased the average steady state weight loss by less than 35% [8]. A further 50% increase from 10 mg to 15 mg of tirzepatide elicited less than a 10% increase in weight loss [8]. Similar diminishing weight loss returns have been observed for semaglutide [17]. Hence, though it has not been carefully tested, the saturating weight loss response to dose frequency that we predict aligns with existing data.

In addition to reducing costs, two additional potential benefits of less frequent dosing are (i) a reduction in side effects and (ii) an increase in persistence and adherence. For (ii), less frequent dosing of GLP-1 products is associated with higher rates of persistence and adherence [11]. While Ref. [11] compare once-daily versus once-weekly doses, their results reflect the general principle that less frequent dosing tends to yield higher adherence [28]. Furthermore, real-world persistence and adherence for GLP-1 products are major impediments to effective therapy. Indeed, in a yearlong study of over 4,000 people, Gleason et al. [11] found that only 1 in 3 persons stayed on their GLP-1 and 27% took their medication as intended. Therefore, significant improvements in real-world efficacy may result from efforts to increase persistence and adherence, including by decreasing the dosing frequency.

As in all mathematical analyses of biomedical systems, our study made a number of simplifying assumptions. For instance, we neglected patient variability in their PK and PD response, though it is known that individual patients vary significantly in their weight loss outcomes from semaglutide [9] and tirzepatide [8]. Our calculations also assumed that patients persist on the medications with perfect adherence (i.e. no discontinuation of treatment and no missed doses), but in fact persistence and adherence are significant problems for actual patients [11, 25]. Understanding how such patient variability and nonadherence affect our predictions presents an important avenue for future research. Furthermore, in light of patient variability, it may be advisable for physicians to try different dose regimens with individual patients to determine the appropriate frequency and dose size required to sustain a desired weight. In fact, dose-dependent side effects may mean that some patients cannot tolerate a dose that is large enough to decrease dose frequency and still sustain their weight loss target. For patients who cannot maintain once weekly dosing (perhaps due to financial constraints), decreasing frequency is likely preferred to simply discontinuing their GLP-1 therapy, since complete discontinuation typically results in regaining two thirds of the lost weight within one year [29]. These limitations notwithstanding, our theory reveals major potential benefits of moving beyond the confines of once-weekly dosing of incretin mimetics, offering significant value for patients, physicians, insurers, and governments.

## Data Availability

The computer code which implements the mathematical model and creates the relevant figures is publicly available at https://github.com/seanlawley/glp1.

## Acknowledgments

SDL and AC were supported by the National Science Foundation (Grant Nos. CAREER DMS-1944574 and DMS-2325258).

