## Supplementary Appendix for "Alternative dosing regimens of GLP-1 receptor agonists may reduce costs and maintain weight loss efficacy"

November 27, 2024

**S.2 BMI and mortality calculations** **7**

*Department of Mathematics, University of Utah, Salt Lake City, UT 84112 USA.

†Tono Health, 90 Furman St. N801, Brooklyn, NY 11201, United States

‡Department of Mathematics, University of Utah, Salt Lake City, UT 84112 USA.

This Supplementary Appendix details the methods used in the main text. The computer code which implements the model and creates the relevant figures is publicly available at `https://github.com/seanlawley/glp1`.

### S.1 PKPD models

We now present the pharmacokinetic (PK) and pharmacodynamic (PD) models of semaglutide and tirzepatide used in this paper. To briefly summarize, our semaglutide PK and PD models are from Strathe et al. [1]. The semaglutide parameters for our "average" virtual patient are taken from Table S2A for PK and Table S3 for PD in the supplementary information of Ref. [1] (except for certain "$E_{\max}$" values, see section S.1.3.1 below). We model tirzepatide PK with the model and parameters of [2], and we use the data of Jastreboff et al. [3] to reparameterize the semaglutide PD model of [1] to model tizepatide PD.

#### S.1.1 Semaglutide PK

Following [1], we model the semaglutide concentration at time $t \geq 0$, denoted $C(t)$, using a one-compartment model,

$$\frac{\mathrm{d}C}{\mathrm{d}t} = -k_{\mathrm{e}}C + k_{\mathrm{a}}\frac{DF}{V}\sum_{n=0}^{\lfloor t/\tau \rfloor} e^{-k_{\mathrm{a}}(t-n\tau)}, \tag{S.1}$$

where $k_{\mathrm{a}}$ and $k_{\mathrm{e}}$ are the respective first-order absorption and elimination rates, $D$ is the dose size, $F \in (0,1]$ is the bioavailability, $V$ is the volume of distribution, $\tau$ is the dosing interval, and $\lfloor t/\tau \rfloor$ denotes the largest integer less than or equal to $t/\tau$. The semaglutide PK parameter values from [1] are given in Table S.1.

#### S.1.2 Tirzepatide PK

Following [2], we model the tirzepatide concentration using a two-compartment model,

$$\begin{aligned} \frac{\mathrm{d}C}{\mathrm{d}t} &= -k_{12}C + k_{21}C_{\mathrm{per}} - k_{\mathrm{e}}C + k_{\mathrm{a}}\frac{DF}{V}\sum_{n=0}^{\lfloor t/\tau \rfloor} e^{-k_{\mathrm{a}}(t-n\tau)}, \\ \frac{\mathrm{d}C_{\mathrm{per}}}{\mathrm{d}t} &= k_{12}C - k_{21}C_{\mathrm{per}}, \end{aligned}$$

where $C(t)$ and $C_{\mathrm{per}}$ denote the respective concentrations at time $t \geq 0$ in the central and peripheral compartments, $k_{12}$ denotes the transfer rate from the central to peripheral compartment, $k_{21}$ denotes the transfer rate from the peripheral to central compartment, and $k_{\mathrm{a}}$, $k_{\mathrm{e}}$, $D$, $F$, $V$, and $\tau$ are as in section S.1.1. The tirzepatide PK parameter values from [2] are given in Table S.1.

##### S.1.3 Semaglutide PD

Our semaglutide PD model and parameters are taken directly from [1]. The one modification is that we use a time-dependent drug concentration rather than the weekly average concentration used in [1], but we show in section S.1.5 below that this choice negligibly affects our results.

The body weight at time $t \geq 0$ after the start of treatment is

$$W(t) = W_0(1 - E_{\mathrm{pl}}(t) - E_{\mathrm{fast}}(t) - E_{\mathrm{slow}}(t)),$$

where $W_0$ denotes the body weight at the start of treatment and $E_{\mathrm{pl}}(t)$, $E_{\mathrm{fast}}(t)$, and $E_{\mathrm{slow}}(t)$ denote the dimensionless placebo, fast, and slow drug effects, respectively. The percentage change in body weight from the drug is thus equal to

$$\frac{W(t) - W_0}{W_0} = -\big(E_{\mathrm{pl}}(t) + E_{\mathrm{fast}}(t) + E_{\mathrm{slow}}(t)\big).$$

Following Strathe et al. [1], the placebo effect is the sum of a transient placebo effect and a slow placebo effect,

$$E_{\mathrm{pl}}(t) = E_{\mathrm{pl,tr}}(t) + E_{\mathrm{pl,slow}}(t),$$

which evolve according to the following ordinary differential equations (ODEs),

$$\begin{aligned} \frac{\mathrm{d}E_{\mathrm{pl,tr}}}{\mathrm{d}t} &= k_{\mathrm{pl,tr}}(K_{\mathrm{pl,tr}} e^{-\gamma_{\mathrm{pl,tr}} t} - E_{\mathrm{pl,tr}}), \\ \frac{\mathrm{d}E_{\mathrm{pl,slow}}}{\mathrm{d}t} &= k_{\mathrm{pl,slow}}(E_{\mathrm{pl,slow},\infty} - E_{\mathrm{pl,slow}}). \end{aligned}$$

We note that we were able to obtain similar results by omitting the transient placebo effect (i.e. setting $E_{\mathrm{pl,tr}}(t) = 0$ for all $t$), but we retain the transient placebo effect to be consistent with [1]. The fast and slow drug effects satisfy

$$\begin{aligned} \frac{\mathrm{d}E_{\mathrm{fast}}}{\mathrm{d}t} &= k_{\mathrm{fast}}\Big(\frac{E_{\mathrm{max,fast}} C(t)}{\mathrm{EC}_{50,\mathrm{fast}} + C(t)} - E_{\mathrm{fast}}\Big), \\ \frac{\mathrm{d}E_{\mathrm{slow}}}{\mathrm{d}t} &= k_{\mathrm{slow}}\Big(\frac{E_{\mathrm{max,slow}} C(t)}{\mathrm{EC}_{50,\mathrm{slow}} + C(t)} - E_{\mathrm{slow}}\Big), \end{aligned}$$

where $C(t)$ is the drug concentration obtained from the PK model in section S.1.1 (the drug mass is converted to moles using the molar mass $\mu$). Following [1], we set $k_{\mathrm{out,fast}} = \infty$ so that $E_{\mathrm{fast}}$ is given by

$$E_{\mathrm{fast}}(t) = \frac{E_{\mathrm{max,fast}} C(t)}{\mathrm{EC}_{50,\mathrm{fast}} + C(t)}.$$

It is straightforward to solve these ODEs to obtain

$$\begin{aligned} E_{\mathrm{pl,tr}}(t) &= K_{\mathrm{pl,tr}} k_{\mathrm{pl,tr}} \int_0^t e^{-k_{\mathrm{pl,tr}}(t-s)} e^{-\gamma_{\mathrm{pl,tr}} s} \, \mathrm{d}s, \\ E_{\mathrm{pl,slow}}(t) &= E_{\mathrm{pl,slow},\infty}(1 - e^{-k_{\mathrm{pl,slow}} t}), \\ E_{\mathrm{slow}}(t) &= k_{\mathrm{slow}} \int_0^t e^{-k_{\mathrm{pl,tr}}(t-s)} \frac{E_{\mathrm{max,slow}} C(s)}{\mathrm{EC}_{50,\mathrm{slow}} + C(s)} \, \mathrm{d}s, \end{aligned}$$

| Symbol | Description | Semaglutide | Tirzepatide |
|---|---|---|---|
| $D$ | dose amount | 2.4 mg [1] | 2.5–15 mg [3] |
| $k_{\mathrm{a}}$ | PK absorption rate | 4.973/wk [1] | 6.266/wk [2] |
| $k_{\mathrm{e}}$ | PK elimination rate | 0.678/wk [1] | 3.090/wk [2]* |
| $V/F$ | apparent volume of distribution | 12.1 L [1] | 3.08 L [2] |
| $\tau$ | dosing interval | varied | varied |
| $k_{12}$ | central to peripheral rate | - | 11.836/wk [2]* |
| $k_{21}$ | peripheral to central rate | - | 7.345/wk [2]* |
| $\mu$ | molar mass | 4114/gmol [4] | 4814/gmol [5] |
| $k_{\mathrm{pl,tr}}$ | rate for transient placebo effect | 0.00885/wk [1] | 0.00620/wk |
| $K_{\mathrm{pl,tr}}$ | placebo effect magnitude | 34.4% [1] | 34.1% |
| $\gamma_{\mathrm{pl,tr}}$ | transient placebo decay rate | 0.0794/wk [1] | 0.2702/wk |
| $k_{\mathrm{pl,slow}}$ | rate for slow placebo effect | 0.0319/wk [1] | 0.0975/wk |
| $E_{\mathrm{pl,slow},\infty}$ | steady-state slow placebo effect | 1% [1] | 2.19% |
| $E_{\mathrm{max,fast}}$ | maximum fast effect | 4.19%** | 2.59% |
| $\mathrm{EC}_{50,\mathrm{fast}}$ | half-max. conc. for fast effect | 48 nM [1] | 45.68 nM |
| $k_{\mathrm{slow}}$ | rate for slow effect | 0.0319/wk [1] | 0.0364/wk |
| $E_{\mathrm{max,slow}}$ | maximum slow effect | 22.6%** | 27.6% |
| $\mathrm{EC}_{50,\mathrm{slow}}$ | half-max. conc. for slow effect | 48 nM [1] | 112.89 nM |

Table S.1: Semaglutide and tirzepatide model parameters. The first 8 rows give PK parameters and the final 10 rows give PD parameters. Parameter values with an asterisk (*) were scaled allometrically [6], meaning that the parameter value reported in [2] was multiplied by the factor $(104.8/70)^{0.8}$, which converts a parameter for a 70 kg body weight [2] to one for a 104.8 kg body weight [3]. Tirzepatide PD parameters were estimated in this study. See section S.1.3.1 for details on the values of $E_{\mathrm{max,fast}}$ and $E_{\mathrm{max,slow}}$ for semaglutide (marked by **).

where we have assumed zero initial conditions,

$$E_{\mathrm{pl,tr}}(0) = E_{\mathrm{pl,slow}}(0) = E_{\mathrm{fast}}(0) = E_{\mathrm{slow}}(0) = 0,$$

which reflects that the therapy starts at time $t = 0$. Parameter values are given in Table S.1.

###### S.1.3.1 Semaglutide $E_{\max}$ values

The parameters $E_{\mathrm{max,fast}}$ and $E_{\mathrm{max,slow}}$ were not reported in Strathe et al. [1] for a single "average" patient. Rather, the following values are reported for one specific patient demographic and health profile,

$$E_{\mathrm{max,fast}} = 3.99\%, \quad E_{\mathrm{max,slow}} = 26.2\%,$$

and then it was specified how these parameters vary for different demographic or health characteristics.

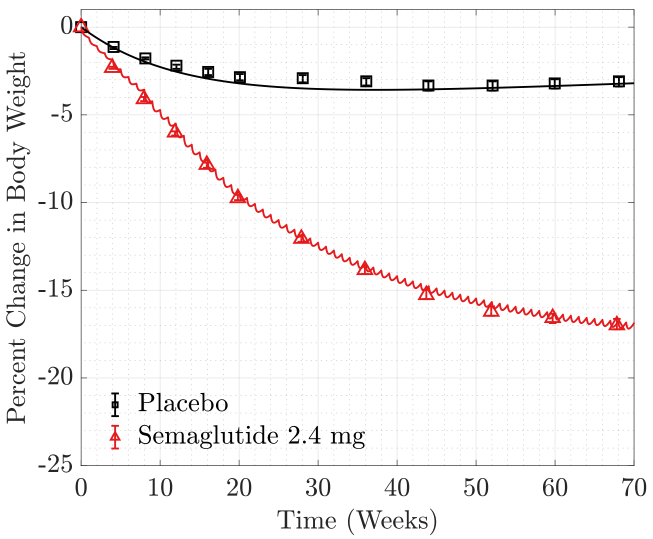

Figure S.1: Solid curves are the semaglutide PD model. Markers are clinical trial data from Wilding et al. [7].

For our model, we choose the following parameter values,

$$E_{\text{max,fast}} = 4.19\%, \quad E_{\text{max,slow}} = 22.6\%. \tag{S.2}$$

We chose the values in Equation (S.2) to fit data from Wilding et al. [7], which was also fit by Strathe et al. [1] (see Figure 2A in [1]). Our resulting fit is shown in Figure S.1. The values in Equation (S.2) thus describe an average patient without any prior knowledge of their demographics (i.e. age, sex, etc.).

To show that our main conclusions are insensitive to the specific values of $E_{\text{max,fast}}$ and $E_{\text{max,slow}}$, in Figure S.2 we plot the weight loss efficacy relative to once-weekly dosing against the cost relative to once-weekly dosing for less frequent dosing (in particular, for dosing intervals of length 7, 8, ..., 28 days). This plot assumes that cost is proportional to the dose frequency. This plot shows these curves for letting $E_{\text{max,fast}}$ and $E_{\text{max,slow}}$ vary from the values in Equation S.2. In particular, the various multicolored curves in Figure S.2 correspond to setting

$$E_{\text{max,fast}} = \phi\, 4.19\%, \quad E_{\text{max,slow}} = \psi\, 22.6\%,$$

where the factors $\phi$ and $\psi$ take the following 11 different values,

$$\phi \in \{0.5, 0.6, \dots, 1.4, 1.5\}, \quad \psi \in \{0.5, 0.6, \dots, 1.4, 1.5\}.$$

The key feature of Figure S.2 is that the $11 \times 11 = 121$ multicolored solid curves are very near each other. Hence, our main conclusion that reducing dose frequency does not commensurately reduce weight loss efficacy holds regardless

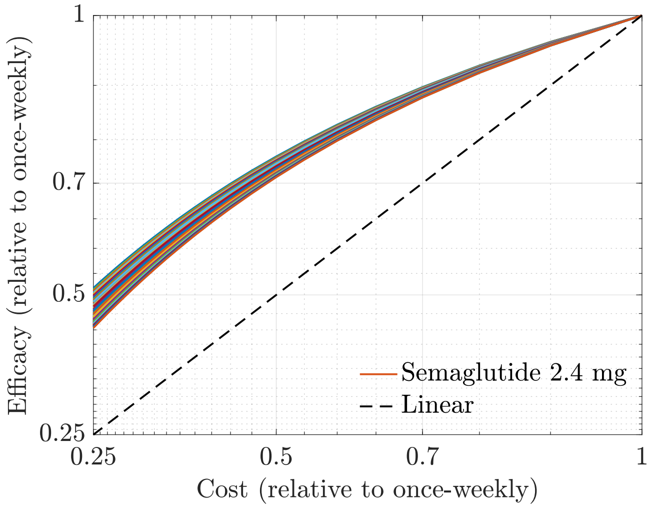

Figure S.2: Reducing cost by reducing dose frequency does not commensurately reduce weight loss efficacy for a wide range of values of the parameters $E_{\text{max,fast}}$ and $E_{\text{max,slow}}$. See section S.1.3.1 for details.

of the exact values of $E_{\text{max,fast}}$ and $E_{\text{max,slow}}$. For instance, notice that all the multicolored curves in Figure S.2 predict that a patient who switches from a 7 day dosing interval to a 14 day dosing interval maintains between 71% to 75% of their weight loss, even though their relative dosing frequency (and thus cost) is cut in half.

##### S.1.4 Tirzepatide PD

We model the PD of tirzepatide using the semaglutide PD model in section S.1.3 after reparameterizing the model to fit the data in [3]. The drug concentration $C(t)$ used in the tirzepatide PD model is obtained from the PK model in section S.1.2.

##### S.1.5 Semaglutide average versus time-dependent concentration

The semaglutide PKPD model that we employ in this paper (detailed above in sections S.1.1 and S.1.3) is identical to the model of Strathe et at. [1], except for one minor modification. In our model, the time-dependent semaglutide concentration, $C(t)$, is used in the PD model. In [1], the semaglutide concentration used in the PD model is an average of the prior week (which we denote by $C_{\text{avg}}$). However, this minor modification only negligibly affects the PD model. We demonstrate this point in Figure S.3, which shows the percent change in

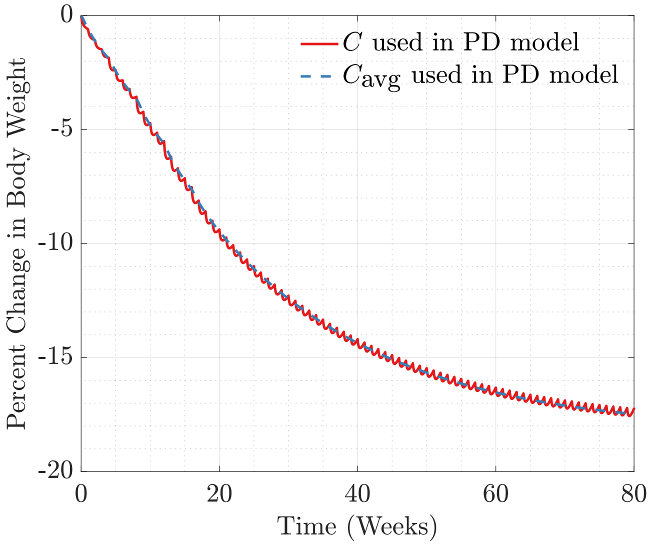

Figure S.3: Percent change in body weight over time for 2.4 mg once weekly dose of semaglutide. The curves are the semaglutide model in section S.1.3 using either the time-dependent semaglutide concentration $C$ (red solid curve) or the weekly averaged semaglutide concentration $C_{\text{avg}}$ (blue dashed curve).

body weight for each version of the model. Indeed, other than small oscillations, the two curves are nearly indistinguishable.

#### S.2 BMI and mortality calculations

To obtain BMI estimates, we first estimated the current US adult BMI distribution from Figure 2 in [8]. Let $p(x)$ denote the probability density function of this current US adult BMI distribution. If a fraction $f$ of obese US adults reduces their BMI by a factor of $a$ (where obesity means a BMI of at least 30 kg/m$^2$), then the resulting BMI distribution has probability density function

$$\overline{p}(x) = \begin{cases} p_0(x) & \text{if } x < 30a, \\ p_0(x) + (f/a)p_0(x/a) & \text{if } 30a \leq x < 30, \\ (1-f)p_0(x) + (f/a)p_0(x/a) & \text{if } x \geq 30. \end{cases} \qquad \text{(S.3)}$$

Here $x$ measures BMI in units of kg/m$^2$ and $0 \leq f \leq 1$ and $0 < a < 1$. To understand (S.3) in words, notice that the distribution is unchanged if $x < 30a$, since (i) adults with BMI less than 30 do not have their BMI reduced (i.e. the weight-loss treatment is only applied to adults with BMI at least 30) and (ii) the treatment cannot cause a person to have BMI less than 30a. Further, the number of adults with BMI between $30a$ and 30 increases because some obese adults decreased their BMI to a level between $30a$ and 30. Finally, for a BMI $x \geq 30$, the distribution changes due to both (a) a decrease since some adults who had a BMI of $x$ reduced their BMI and (b) an increase since some adults with a larger BMI (namely, $x/a$) reduced their BMI to $x$.

The fraction $f_1$ of obese US adults that can receive q1wk dosing for a given annual supply of semaglutide was computed according to

$$f_1 = \frac{\text{(mg of semaglutide annual supply)}}{\text{(2.4 mg per week)(52 weeks)(number of obese US adults)}},$$

where the number of obese US adults was set to $1.066 \times 10^8$ [8]. The fraction $f_2$ that can receive q2wk dosing was set to $f_2 = 2f_1$ where $0 < f_1 \leq 0.5$. The results in the paper were computed from (S.3) with either $a = 1 - 0.19 = 0.81$ and $f = f_1$ for q1wk dosing or $a = 1 - 0.14 = 0.86$ and $f = f_2 = 2f_1$ for q2wk dosing.

To estimate the number of lives saved, we first estimated the number of annual deaths by multiplying the number of adults in each of the standard seven BMI categories (using the standard BMI cutoffs between categories of 18.5, 25, 27.5, 30, 35, and 40) by the BMI category-dependent annual mortality rates reported in [8]. The number of lives saved was then obtained by comparing each hypothetical scenario (i.e. for each distribution $\overline{p}$ in (S.3)) to the current scenario.

[4] National Center for Biotechnology Information. Pubchem compound summary for CID56843331, semaglutide, 2024. Retrieved September 17, 2024.

[5] National Center for Biotechnology Information. Pubchem compound summary for CID11977456, tirzepatide, 2024. Retrieved September 17, 2024.

[6] European Medicines Agency. EPAR assessment report: Mounjaro (h-c-005620-ii-0007), 2024. Accessed: November 6, 2024.

[7] John PH Wilding, Rachel L Batterham, Salvatore Calanna, Melanie Davies, Luc F Van Gaal, Ildiko Lingvay, Barbara M McGowan, Julio Rosenstock, Marie TD Tran, Thomas A Wadden, et al. Once-weekly semaglutide in adults with overweight or obesity. *New England Journal of Medicine*, 384(11):989–1002, 2021.

[8] Abhishek Pandey, Yang Ye, Chad R Wells, Burton H Singer, and Alison P Galvani. Estimating the lives that could be saved by expanded access to weight-loss drugs. *Proceedings of the National Academy of Sciences*, 121(43):e2412872121, 2024.
